# Population age structure only partially explains the large number of COVID-19 deaths at the oldest ages

**DOI:** 10.1101/2020.04.09.20056341

**Authors:** Anthony Medford, Sergi Trias-Llimós

## Abstract

To date any attention paid to the age shape of COVID-19 deaths has been mostly in relation to attempts to understand the differences in case fatality rates between countries. The aim of this paper is to explore differences in age distribution of deaths from COVID-19 among European countries which have old age structures. We do this by way of a cross-country comparison and put forward some reasons for potential differences.

## Background

On 11, March 2020 the World health Organization declared COVID-19 to be a pandemic. Despite the variation in symptomatology and health complications from the disease presents greater health and mortality risks in individuals at higher ages and in those with pre-existing medical conditions. In the worst affected countries, there are spikes in hospitalizations, ICU admissions and deaths. The health systems of these countries are being stretched to the limit. The morbidity impact of the virus is attracting considerable attention, especially as the availability of intensive care units is being exceeded by the number of cases requiring critical care.

As the pandemic matures the numbers of deaths is rising at an alarming rate. On March 15, 2020 there were about 6,500 deaths globally. Thirteen days later, on March 28, 2020 there were almost 31000 deaths globally -- an almost fivefold increase. Surprisingly, there has not been much focus on the age pattern of COVID-19 deaths thus far though attempts have been made to understand the differences in case fatality rates between countries (1–3).

We explore differences in the age distribution of COVID-19 deaths among European countries with older age structures: France, Italy, the Netherlands and Spain (subsample). We do this via a cross-country comparison of the age-patterns in observed COVID-19 death counts and their counterfactual distribution adjusted by the age-structure of Italy. We include Chinese data for comparison. See Supplementary Material for methodological details.

### Observed deaths distribution

While deaths occur more frequently at the higher ages, there are differences both in the observed age-distribution and in the estimated counterfactual distribution (Figure 1). Deaths in China occur at slightly younger ages than in Europe and are split slightly more evenly across the older ages with 30% of deaths in age bands 60-69 and 70-79 and 20% of deaths at ages above 80. Above age 80, the proportion of deaths is 50%, 58%, 59% and 59% for Italy, the Netherlands, Spain and France respectively. These differences between the countries persist after adjustment for age-structure.

**Figure 1.**
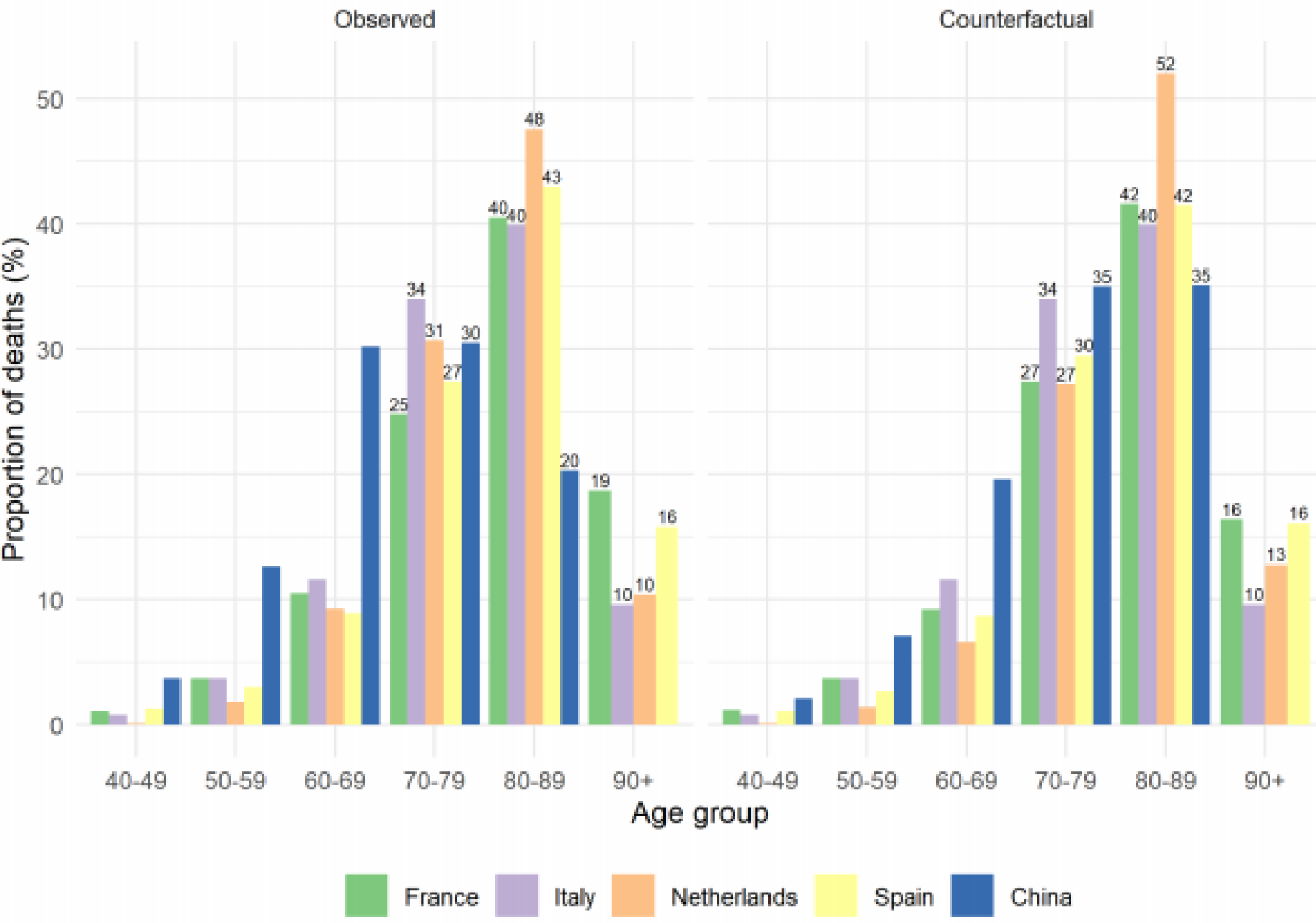
COVID-19 death distribution for Italy, Spain*, France**, the etherlands and China***. Observed and counterfactual adjusting by age-structure of Italy. *Subsample. **As at March 30. ***As at March 22, 2020. All other countries as at March 31, 2020.

## Discussion

Italy has the second oldest population the world (4) and the oldest in Europe but we observe proportionately fewer deaths there from COVID-19 at the older ages (80+ and 90+) than either Spain, France or the Netherlands even though Italy has a larger population at those ages. This phenomenon is particularly evident in the case of the Netherlands where the population difference to Italy at ages above age 80 is in the millions. Therefore, age structure is only partially predictive of the deaths at older ages. Explanations have previously been attributed to variations in the definition of COVID-19 related deaths (1), testing strategies (1) and transmission pathways (3).

The age distribution of deaths gives clues tothe epidemic’s impact on the most affected age groups. However, the estimated age distributions depend critically on data quality and on the comparability of COVID-19 mortality across countries. Presently, information from official sources can be scarce and of variable quality. For example, it appears that non-hospital COVID-19 deaths are not recorded in France and COVID- 19 deaths in Spanish nursing homes are under-recorded (5).

We suspect that there may be some effects related to the predominant model of eldercare delivery within a country. For example, in the Netherlands there is a high number of nursing care homes whereas in Italy and Spain much care is provided at home by children and grandchildren. Second, we could be observing the outworking of country-specific cohort effects where particular cohorts are impacted differently between different countries. Third, differences may be reflective of different levels in old age frailty between countries. The countries with the most robust individuals at a given age group would be less likely to succumb to COVID-19. Finally, underlying medical conditions which are exacerbated by the presence of COVID-19 and viceversa may be more prevalent in some populations compared to others. This would result in more severe illness and ultimately, higher deaths in one country versus the other.

In summary, we note that the countries included in this study are in different stages of the COVID-19 epidemic. We conjecture that as the epidemic evolves the relative number of deaths at age 80 and above may decline. The differences in the age-patterns of COVID-19 deaths are not only dependent on the age-structure of a given population, but also on other country-specific factors. A better grasp of these is crucial to gain a greater understanding of the age pattern of COVID-19 deaths.

## Data Availability

Links to data in manuscript.

## Supplementary Material

| Age | *China |  | Italy |  | Spain |  | Netherlands |  | France |  |
| --- | --- | --- | --- | --- | --- | --- | --- | --- | --- | --- |
|  | count | Dist'n | count | Dist'n | count | Dist'n | count | Dist'n | count | Dist'n |
| **0-9** | 0 | 0% | 0 | 0% | 1 | 0% | 0 | 0% | 0 | 0% |
| **10-19** | 1 | 0% | 0 | 0% | 1 | 0% | 0 | 0% | 1 | 0% |
| **20-29** | 7 | 1% | 2 | 0% | 6 | 0% | 0 | 0% | 3 | 0% |
| **30-39** | 18 | 2% | 25 | 0% | 14 | 0% | 0 | 0% | 13 | 0% |
| **40-49** | 38 | 4% | 92 | 1% | 44 | 1% | 1 | 0% | 33 | 1% |
| **50-59** | 130 | 13% | 409 | 4% | 101 | 3% | 19 | 2% | 111 | 4% |
| **60-69** | 309 | 30% | 1271 | 12% | 301 | 9% | 97 | 9% | 315 | 11% |
| **70-79** | 312 | 30% | 3724 | 34% | 929 | 27% | 319 | 31% | 742 | 25% |
| **80-89** | 208 | 20% | 4366 | 40% | 1461 | 43% | 495 | 48% | 1209 | 40% |
| **90+** |  |  | 1051 | 10% | 538 | 16% | 108 | 10% | 559 | 19% |
| **Total** | 1023 | 100% | 10940 | 100% | 3396 | 100% | 1039 | 100% | 2986 | 100% |

**A.1:** COVID-19 death distribution for Italy, Spain\*, France\*\*, the Netherlands and China\*\*\*.
Observed and counterfactual adjusting by age-structure of Italy. \*Subsample. \*\*As at March 30. \*\*\*As at March 22, 2020. All other countries as at March 31, 2020.

### Data

#### Covid-19 deaths

China: 11 February 2020 https://cdn.onb.it/2020/03/COVID-19.pdf.pdf

Italy: 28 March 2020 https://www.epicentro.iss.it/coronavirus/bollettino/Infografica_28marzo%20ITA.pdf

Spain: 28 March 2020, 21.00. https://www.mscbs.gob.es/en/profesionales/saludPublica/ccayes/alertasActual/nCov-China/documentos/Actualizacion_58_COVID-19.pdf

Netherlands: 31 March 2020, 10.00 https://www.rivm.nl/documenten/epidemiologische-situatie-covid-19-in-nederland-29-maart-2020

France: 22 March 2020, https://dc-covid.site.ined.fr/

#### Population

Human Mortality Database for Italy (2017), Spain (2016) the Netherlands (2016), and France (2017). Data for China (2019) came from https://www.populationpyramid.net/china/2019/

### Methods

Proportion of deaths for each age group was estimated dividing the number of deaths by the total number of deaths in the country.

For each age group *i* we estimated the counter-factual proportion of Covid-19 deaths under the assumption that all countries have the Italian age-specific distribution. Our approach is based on indirect standardization techniques, and we did so by applying the following formula in each of the countries

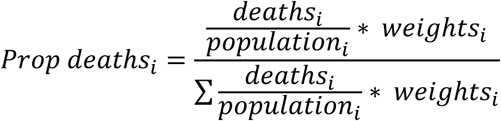

Where weights stand for the proportion of population in age group i in the standard population (Italy), deaths are the Covid-19 observed deaths and population is the population at the country.

